# Prediction of the Epidemic of COVID-19 Based on Quarantined Surveillance in China

**DOI:** 10.1101/2020.02.27.20027169

**Authors:** Rui Li, Wenliang Lu, Xifei Yang, Peihua Feng, Ozarina Muqimova, Xiaoping Chen, Gang Wei

## Abstract

**Backgroud and Objective:** To predict the epidemic of COVID-19 based on quarantined surveillance from real world in China by modified SEIR model different from the previous simply mathematical model.

**Design and Methods:** We forecasted the epidemic of COVID-19 based on current clinical and epidemiological data and built a modified SEIR model to consider both the infectivity during incubation period and the influence on the epidemic from strict quarantined measures.

**Results:** The peak time of the curve for the infected newly diagnosed as COVID-19 should substantially present on Feb. 5, 2020 (in non-Hubei areas) and Feb. 19, 2020 (in Hubei). It is estimated that the peak of the curve of the cumulative confirmed cases will appear in non-Hubei areas on Mar. 3, 2020 and in Hubei province on Mar. 10, 2020, and the total number of the patients diagnosed as COVID-19 is 18,000 in non-Hubei areas and 78,000-96,000 in Hubei. The Chinese COVID-19 epidemic can be completetly controlled in May, 2020.

**Conclusions:** COVID-19 is only a local outbreak in Hubei Province, China. It can be probably avoided the pandemic of global SARS-CoV-2 cases rise with the great efforts by Chinese government and its people.

## Background and Objective

The first case of pneumonia of unknown aetiology was reported in Wuhan, Hubei Province in China on Dec. 8, 2019, and the epidemic spread rapidly in Wuhan, Hubei Province and the surrounding areas of Hubei Province.^**1**, **2**^ A novel coronavirus was identified as the causative agent by the Chinese authorities on Jan. 7, 2020, and on Jan. 10, 2020, the World Health Organization (WHO) designated the novel coronavirus as 2019-nCoV, and later defined the pneumonia as COVID-19 associated with infecting SARS-CoV-2.

According to the statistics of National Health Commission of People’s Republic of China (www.nhc.gov.cn), by at 12:00 on Feb. 17, 2020, a total of 441 individuals, including 137 individuals in Singapore and Japan together, were diagnosed as COVID-19 outside of China, and a total of 70,639 infected cases in mainland China (with a total population of 1400. 05 million), including 58,182 infected cases were diagnosed as COVID-19 associated with SARS-CoV-2 in Hubei Province (with a total population of 59.4 million, accounting for 4.2% of the total population in mainland China), accounting for 82.4% of the total count of the infected individuals in China (**Figure 1**), and including 41,152 cases in Wuhan (with a total population of 11.08 million), accounting for 70.7% of the total number of the infected individuals in Hubei Province (accounting for 18.7% of the total population in Hubei Province). There were 9 cities in Hubei Province, such as Xiaogan (with a total population of 5 million), Huanggang (with a total population of 7 million), Jingzhou, Suizhou, Xiangyang, Ezhou, Huangshi, Jingmen and Yichang, with more than 1,000 infected patients respectively. And outside of Hubei Province, there were 6 provinces, including Guangdong, Henan, Zhejiang, Hunan, Anhui and Jiangxi, with more than 900 infected patients respectively. The total infected patients in these six provinces accounted for 53.4% of the total infected individuals outside of Hubei Province, particularly in Wenzhou and Shenzhen, the mere two cities, with a total of more than 400 infected persons outside of Hubei Province.

**Figure 1:**
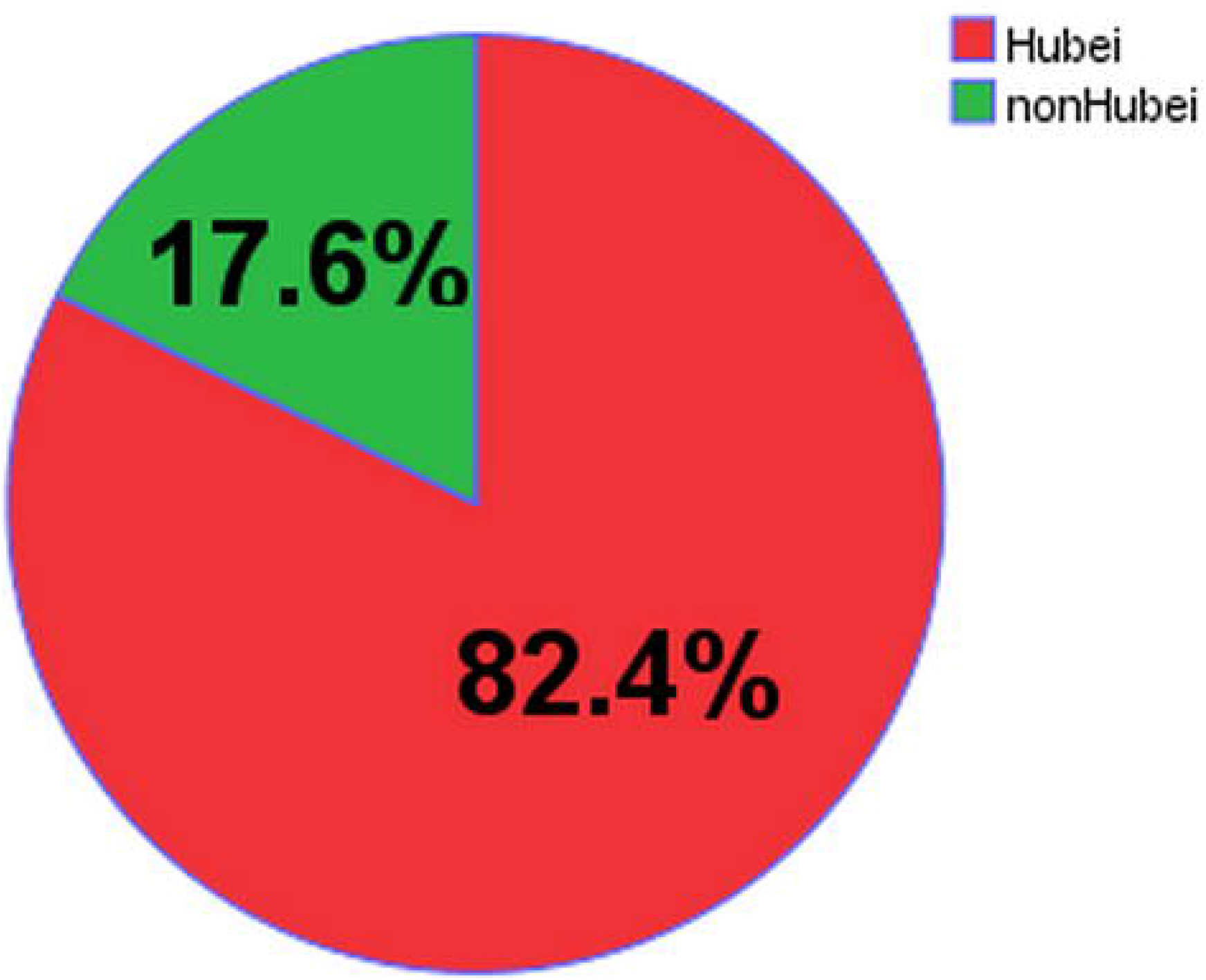
The total count of the infected individuals in Hubei Province accounts for 82.4% of the total number of the infected cases in China although its total population simply accounts for 4.2% of the total population in mainland China.

After Wuhan were sealed off on Jan. 23, 2020, the other cities in Hubei Province successively started to isolate residential quarters and villages and shut off the traffic to halt the population flow in urban and rural regions. According to Tencent’s big data (https://heat.qq.com/) and Location Intelligence (www.wayhe.com), from Jan. 1, 2020 to Jan. 24, 2020, the population outflow from Wuhan is about 5 million, with 65% of the outflow into other cities in Hubei Province, such as Huanggang and Xiaogan (accounting for 27.0% together), and with 55% of the outflow outside of Hubei Province, including Henan, Guangdong, Zhejiang, Hunan, Jiangxi and Anhui together. The remainder population was 9 million in Wuhan, which was the most severe epidemic region. The strict blocked and isolated measures had been taken in other cities in Hubei before the local outbreak of SARS-CoV-2, which had not caused fulminating infection similar to Wuhan. In Hubei and other areas with serious epidemic situation, Chinese government had taken the strict blocked and isolated control, including the infected, the suspiciously infected individuals and the susceptible intimately contacted with the infected and the suspiciously infected individuals, involved the independent community buildings and villages, which were separated and quarantined in a unified way. On Jan. 28, 2020, JAMA magazine interviewed Professor NIH of the United States, and affirmed that the Chinese government adopted the strategy of closure and quarantine against the epidemic.

Many academics tried to estimate key epidemiological characteristics and possible outcomes of the epidemic by means of transmission model mathematically. On Jan. 24, 2020, Jonathan M. Read^**3**^ estimated the basic reproduction index (R0) for SARS-CoV-2 to be between 3.6 and 4.0, then and COVID-19 would outbreak further both in other Chinese cities and in international travel destinations sucn as Thailand, Japan, Taiwan, Hong Kong and South Korea at an sharply increasing rate. According to his model, he predicted the number of infected individuals in Wuhan to be greater than 190 thousand by Feb. 14, 2020 although with a 99% effective reduction as a result of trave restrictions, the size of the epidemic outside of Wuhan might only be reduced by 24.9%. His model suggested that travel limitation from and to Wuhan city were unlikely to be effective in halting transmission across China.

Professor Joseph T Wu^**4**^ published a paper in the Lancet on Jan. 31, 2020. The R0 of SARS-CoV-2 was estimated to be 2.68 by MCMC method, and the SEIR model was used to predict the epidemic.

However, there were some deficiencies in his study:1) the main population outflow from Wuhan was not outside of Hubei Province:his study focused on several large cities (Chongqing, Beijing, Shanghai, Guangzhou and Shenzhen), instead of the moderate-sized urban accounting for 65% in the outflow population from Wuhan;2) the L (T) value referred to the spring transport data in 2019:the comprehensive traffic interdiction in Hubei Province and other severe provinces isolated strictly, after Jan. 23, 2020, the spring transportation passenger flow L (T) should be 0;3) underestimated the strength of the strict isolated measures and the initial and maintaining time by Chinese government:since Jan. 23, both community buildings in cities and villages in rural areas as the separated units, including the unified respective isolation of the infected, the suspiciously infected individuals and the susceptible closely contacted with the infected and the suspiciously infected individuals;4) evaluating R0:there was significant differences between in Hubei and in non-Hubei areas available to the observed data by MCMC model^**5**^5) overestimate the spread scope of the epidemic:from a global perspective, although all continents were scattered, but most of them are imported, and the number of the infected outside of China only accounts for 0.6% together globally by at 12:00 on Feb. 17, 2020; the number of the infected in Hubei accounted for 82.4% of the total number in China, and the number of the infected in Wuhan accounted for 70.7% of the total number in Hubei;6) overestimated the spread of the epidemic:the number of the infected newly diagnosed as COVID-19 in non-Hubei areas had declined steadily for 12 consecutive days since Feb. 4, 2020. In Hubei, because clinical diagnosis had also been considered as the diagnosis criterion (still not announcing the RNA result of SARS-CoV-2) since Feb. 12, most of the suspiciously infected individuals in the inventory were diagnosed as COVID-19 on Feb. 13, and the number of the infected newly diagnosed increased dramatically, but the number of the suspiciously infected individuals reduced by more than 80%. However, the number of the infected newly diagnosed in Hubei had reduced slowly since Feb. 9 in case of the basis of RNA result. From the above, it can be concluded that the epidemic of COVID-19 is still a local public health emergency in Hubei Province of China currently.

The above data from real world in China thew a hit on their predictions on the COVID-19 epidemic simply taking advantage of epidemiological model but ignoring quarantined measures and different R0.

## Design and Methods

There is no quarantined measure in the early stage of the epidemic, so R0, as the basic reproductive index, is often high for SARS-CoV-2,^**6**, **7**^ and there is a significant difference between in Hubei and in non-Hubei areas, but both decrease gradually with the strengthening of isolated measures.^**8**, **9**^

We forecasted the epidemic of COVID-19 by modified SEIR model^**10**, **11**^ and current clinical and epidemiological data on COVID-19, involved in two practical factors to correct the model:1) considering the infectivity during incubation period^**12**^, 2) considering the influence on the epidemic from strict quarantined measures. It had been confirmed that both the latent and the infected are infectious. We assumed that both were with the same infectivity. According to the epidemic data before Feb. 1, 2020, it indicated that the probability from the exposed to the infected was 1.7%,^**13**, **14**^ and the average incubation period was 7 days, and the average hospital stay was 14 days.^**15**, **16**^ The average daily contact rate of the infected was 6, referring to the statistical data from CCDC in the early stage of SARS epidemic in 2003.^**17**, **18**, **19**^ However, the population density in Hubei Province was 2.3 times the national average, so the daily contact rate in Hubei was 13.8, and the average daily contact rate of the latent was twice as the infected. The calculation method was R0 = average daily contact rate× probability of the exposed to the infected ×infection period. The average infection period was 21 days. Therefore, R0 was 4.9-9.9 in Hubei, but 2.5-4.8 in non-Hubei areas. However, R0 would gradually decrease with the progress of the epidemic for the sake of strengthened isolation and blocking measures.^**20**^ The median value of R0 was 7.5 in Hubei. According to the statistical data from Hubei Provincial Center for Disease Control and Prevention (http://wjw.hubei.gov.cn/), there were 27 patients diagnosed as COVID-19 and 0 death in Wuhan on Dec. 31, 2019. Since nobody approached to obtain the quantity of the latents during epidemic period early,^**21**^ we had to deduce the quantity of the latents by the subsequent epidemiological data, the probability from the latent to the infected, which was 14.0%. Consequently, the possible quantity of the latents was 208 individuals in Hubei on Dec. 31, 2019. Assuming that there was only one hidden infected individual with COVID-19 in non-Hubei area at that time, the quantity of the latents was 8 individuals in non-Hubei areas. We forecasted the epidemic of COVID-19 both in Hubei and in non-Hubei areas by modified SEIR model^**22**^ by the software K-SEIR simulator (v1.0), combined with the epidemic data of the last 45 days, and aimed to obtain the possible total number of the infected individuals diagnosed as COVID-19 in Hubei and in non-Hubei areas and the time elimilating the epidemic of SARS-CoV-2 in Hubei and in non-Hubei areas. The modified SEIR model was as listed below (**Table 1**).

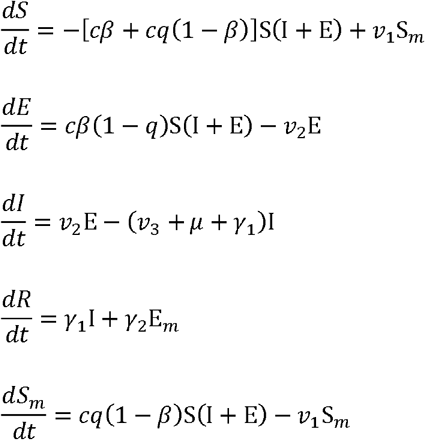

S, E, I, R, S_m,_ E_m_ represent the susceptible individuals, exposed individuals, infected individuals, convalescent individuals, susceptible individuals quarantined for being observed medically and the exposed individuals quarantined respectively. The convertion speed rate of E, S_m,_ E_m_ is *cβ* (1−*q*), *cq*(1−*β*) and *cq* respectively.

**Table 1.** Parameters estimate for COVID-19 in China according to MCMC and CCDC

| Parameter | Definition | Mean value |
| --- | --- | --- |
| c | Contact rate | 13.8 |
| $\beta$ | Probability of transmission per contact | $1.7 \times 10^{-9}$ |
| q | Quarantined rate of the exposed individuals | $5 \times 10^{-7}$ |
| $\nu_1$ | Rate of removing the quarantined( observed medically) who are not infected but intimate contact with the exposed or the infected | 1/14(0.07) |
| $\nu_2$ | Conversion speed rate of the exposed persons to the infected persons | 1/7(0.14) |
| $\nu_3$ | Speed rate of quarantine for the infected individuals | 0.13 |
| $\mu$ | Speed rate of death induced by SARS-CoV-2 | $3.7 \times 10^{-4}$ |
| $\gamma_1$ | Speed rate of recovery for the symptomatic infected individuals | 0.24 |
| $\gamma_2$ | Speed rate of recovery for the asymptomatic infected individuals | 0.30 |

**MCMC**:Markov chain Monte Carlo;**CCDC**:Chinese Center for Disease Control and Prevention

## Results

The quarantined efficiency depended on the occasion and strength of blocking and isolation.^**23**, **24**^ The strength of blocking and isolation included the intensity and scope of containment (**Figure 2**, **Figure 3**). Theoretically, the maximal reducing number of the possible infected individuals could be calculated by the formula:1 - 1/R (1 - 1/7.5) × 100%,^**25**^ about 86.7%. In China, in the early stage of COVID-19 epidemic, the most severe isolated measures had been taken in the areas where epidemic was likely to be serious according to the population outflow from Wuhan. Both the occasion and intensity of quarantined measures were far greater than that during influenza outbreak in the United States in 1918. In this study, it is proved to be 90% efficacy in halting transmission by the blocked and isolated measures, so the size of the epidemic could be reduced by 87.0% × 90%, about 78.0%.

**Figure 2:**
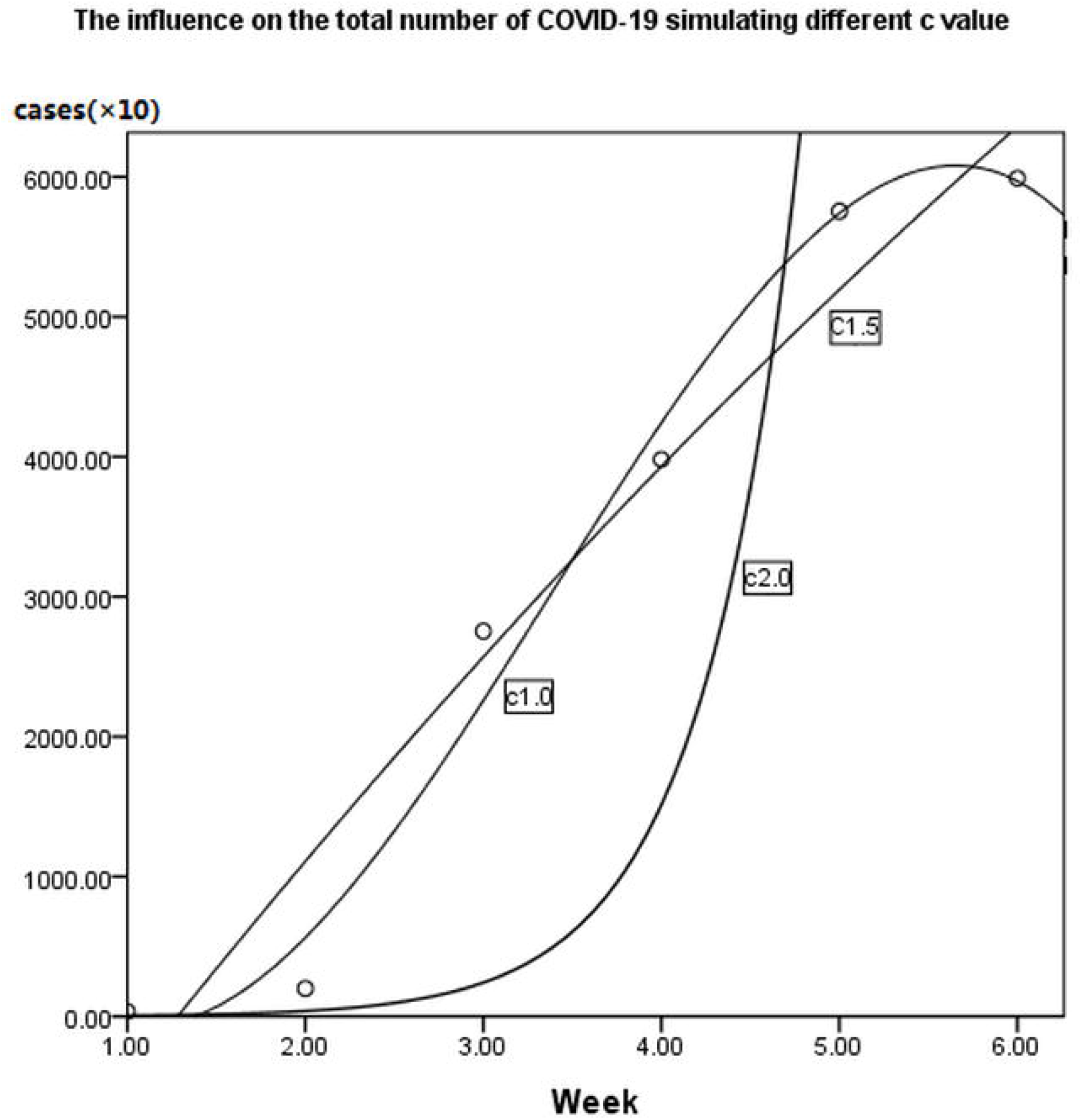
Contact rate *c* indicates the intensity of containment. The figure clarifies the influence on the total count of COVID-19 simulating the different *c* value (1.0×*c*, 1.5×*c*, 2.0×*c*), and the lower *c* value signifies stern surveillance and better halting transmission.

**Figure 3:**
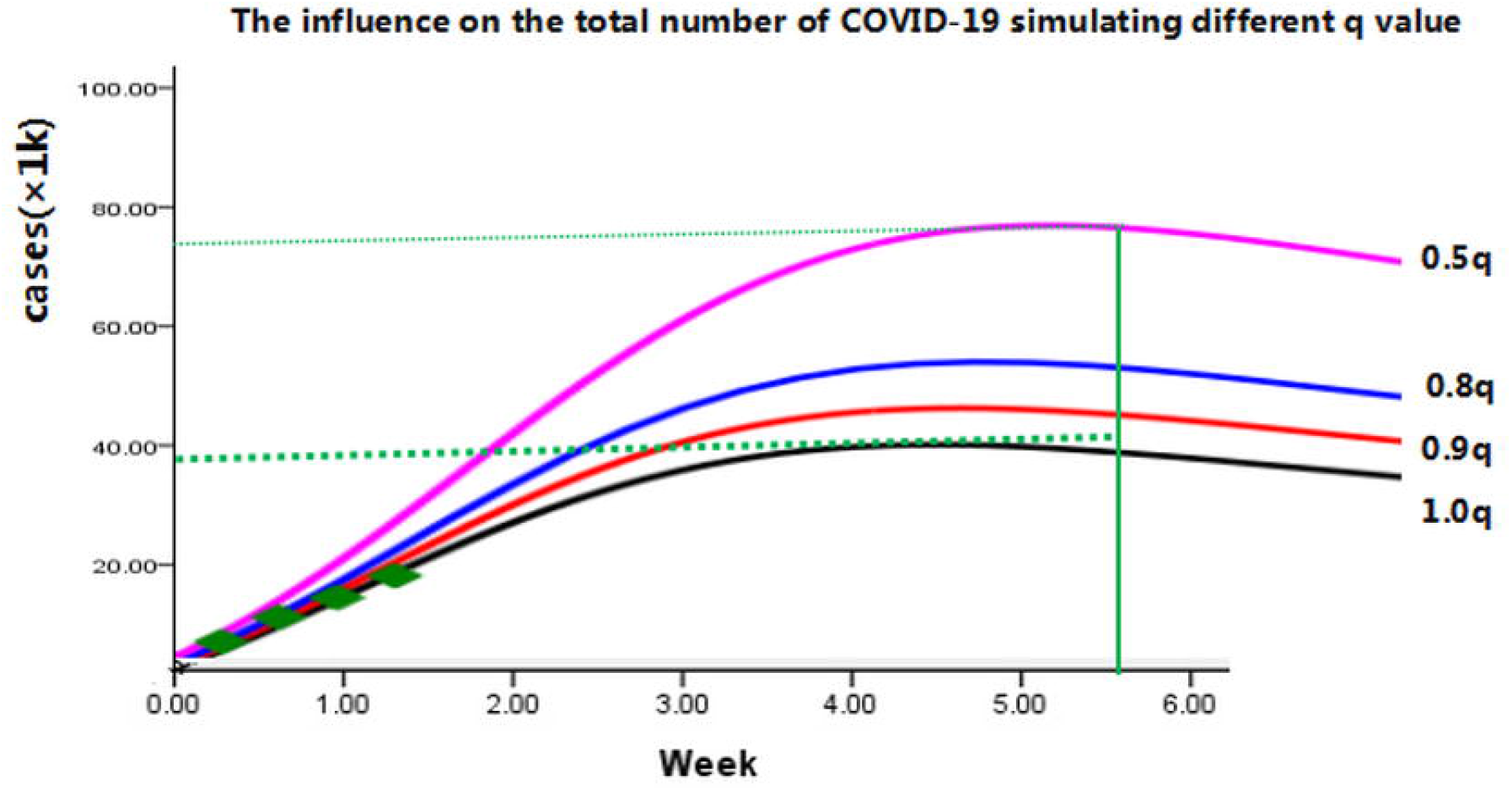
Quarantined rate of the exposed individuals *q* implies the scope of containment. The figure clarifies the influence on the total count of COVID-19 simulating the different *q* value (1.0×*q*, 0.9×*q*, 0.8×*q*, 0.5×*q*), and the lower *q* value (0.5*q*) indicates deficient quarantine on the infected and exposed individuals gives rise to doubling nearly in the total number of COVID-19 compared with 1.0q.

The ultimate population outflow from Wuhan on Jan. 23, 2020 was the most source of transmission, based on the incubation period of 1-14 days, so the peak time of the curve for the infected newly diagnosed as COVID-19 should substantially present on Feb. 5, 2020 (in non-Hubei areas) and Feb. 19, 2020 (in Hubei) for the intensive blocked and isolated measures resulting in population mobility accessible to zero during the Spring Festival (**Figure 4**, **Figure 6**). It is estimated that the peak of the curve of the cumulative confirmed cases will appear in non-Hubei areas on Mar. 3, 2020 and in Hubei province on Mar. 10, 2020, and the total number of the patients diagnosed as COVID-19 is 18,000 in non-Hubei areas and 78,000-96,000 in Hubei. The Chinese COVID-19 epidemic can be completetly controlled in May, 2020 (**Figure 5**, **Figure 7**). It can be probably avoided the pandemic of global SARS-CoV-2 cases rise with the great efforts by Chinese government and its people.^**26**, **27**^

**Figure 4:**
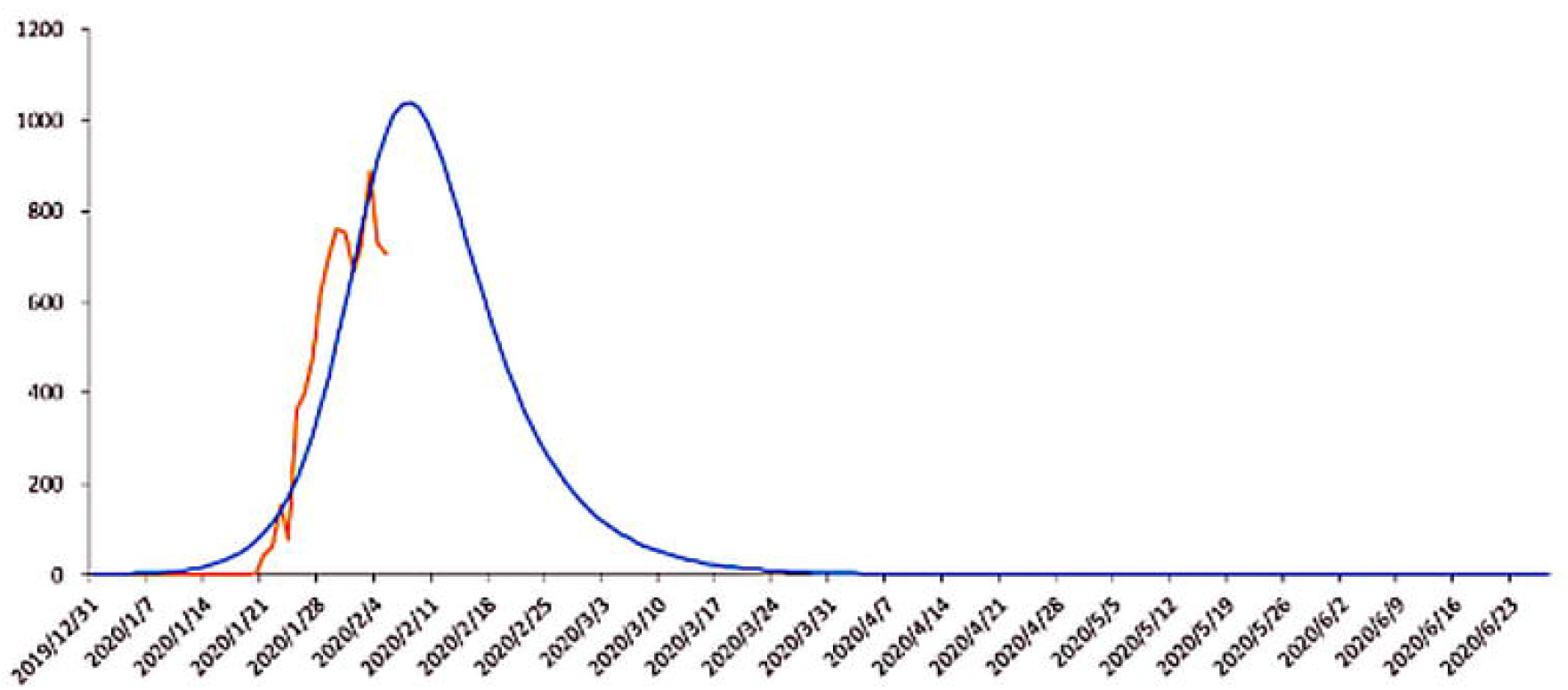
The peak time of the curve for the cases newly diagnosed as COVID-19 should substantially present on Feb. 5, 2020 in non-Hubei areas.

**Figure 5:**
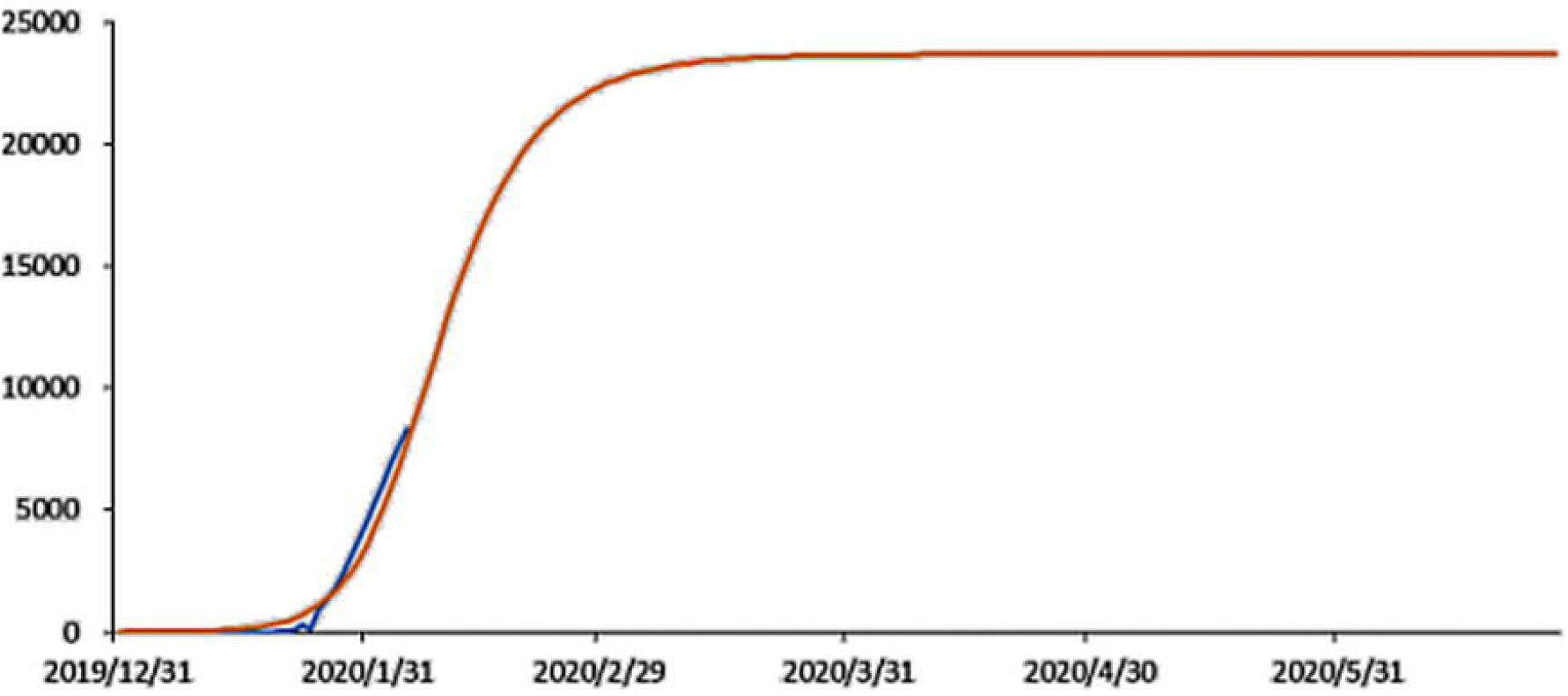
The peak of the curve of the cumulative confirmed cases will appear in non-Hubei areas on Mar. 3, 2020 and the total number of the cases diagnosed as COVID-19 is approximately 18,000 in non-Hubei areas.

**Figure 6:**
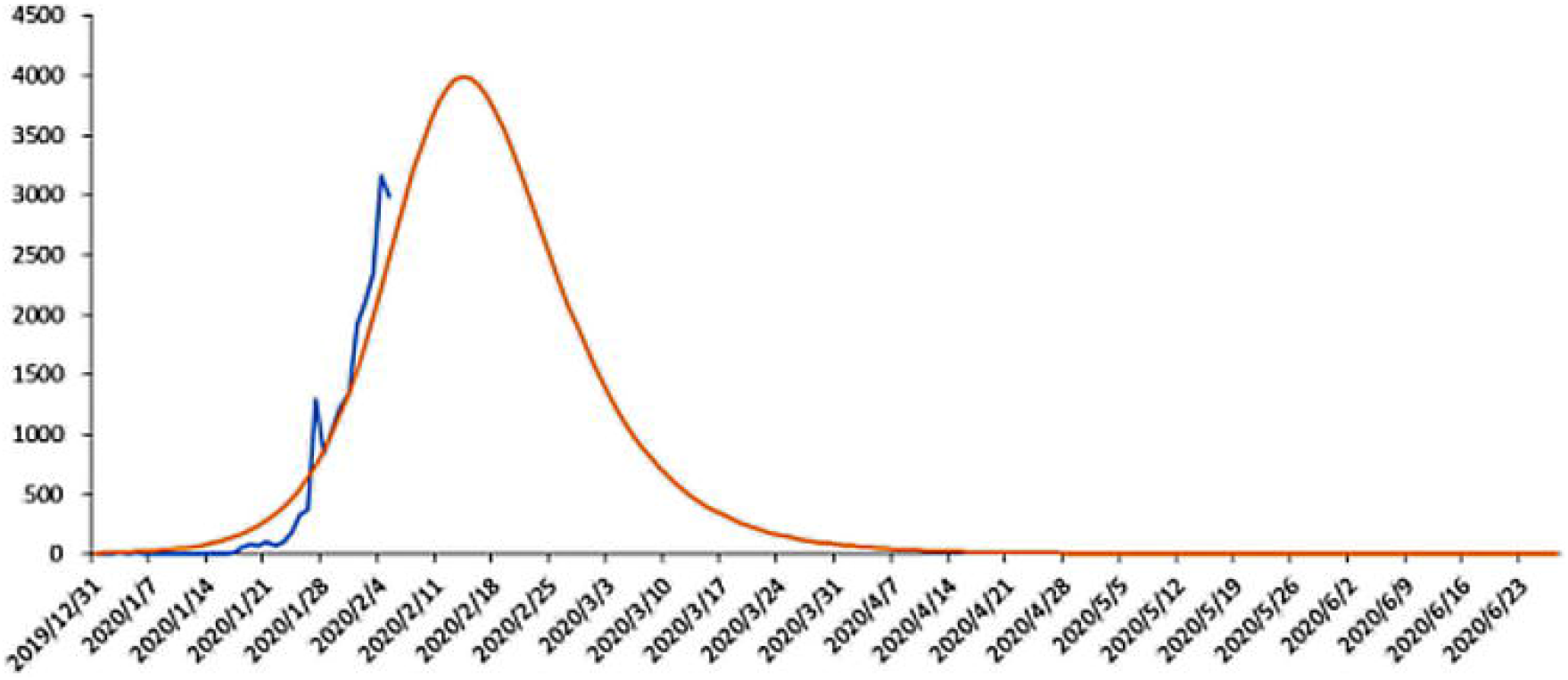
The peak time of the curve for the cases newly diagnosed as COVID-19 should substantially present on Feb. 19, 2020 in Hubei.

**Figure 7:**
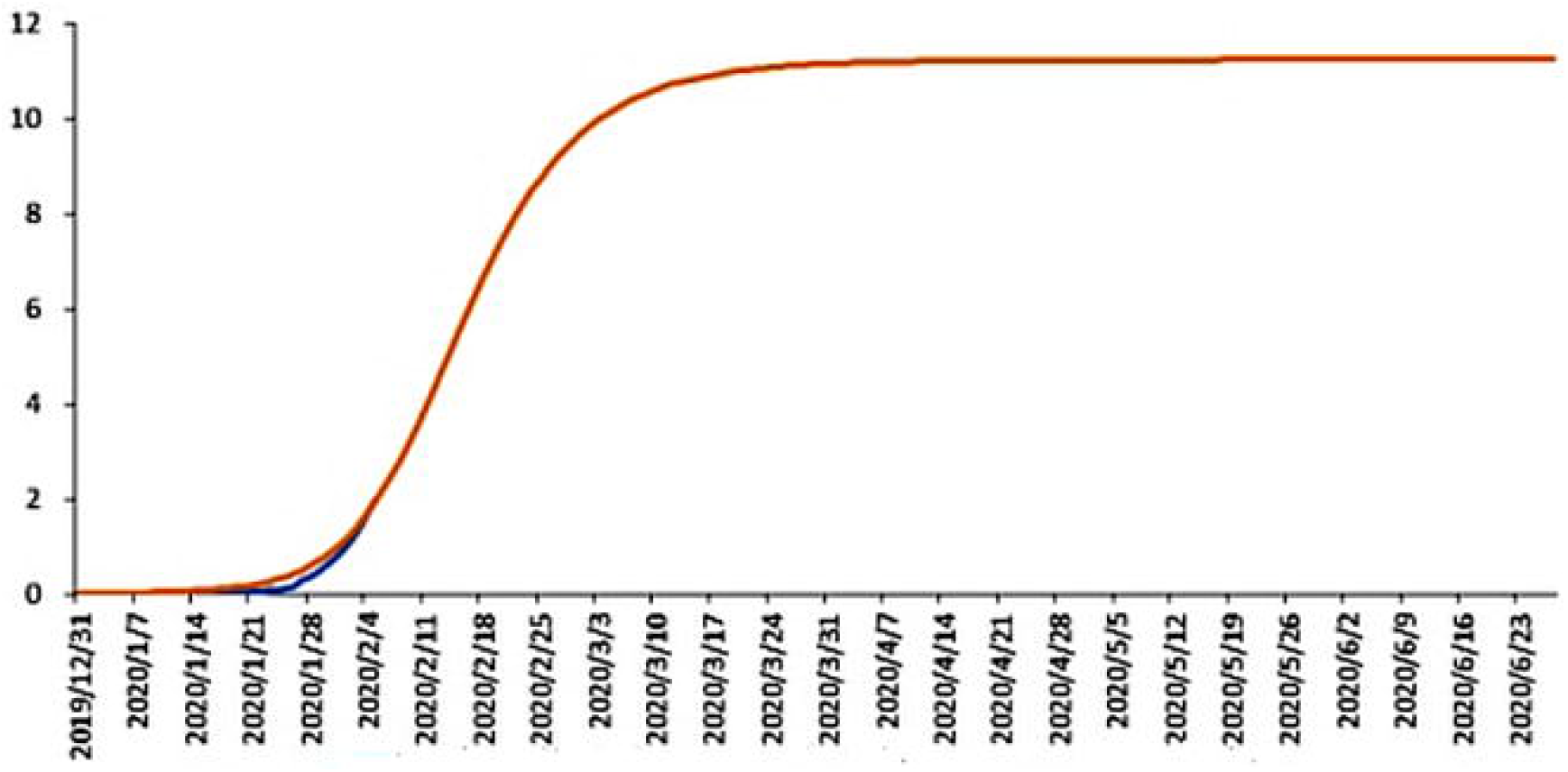
The peak of the curve of the cumulative confirmed cases will appear on Mar. 10, 2020, and the total number of the cases diagnosed as COVID-19 is about 78,000-96,000 in Hubei.

## Discussion and Conclusions

From the above, it could be seen that our study was principally based on the epidemiological characteristics of the COVID-19 in recent 45 days and modified SEIR model, which was quite different from other absolutely theoretical models, mainly as follows:1) our study was based on the epidemiological characteristics of the COVID-19 according to epidemic situation, considering that the incubation period was still infectious;2) evaluating R0 was on the basis of the big data of epidemic situation, R0 was 4.9-9.9 (the median R0 7.5) in Hubei, but 2.5-4.8 (the median R0 3.7) in non-Hubei areas;3) the occasion and intensity of the blocked and isolated measures by Chinese government were unprecedented and efficient in scale, which prevented SARS-CoV-2 outbreaking in non-Hubei area;4) in scale, the COVID-19 epidemic was currently a local public health emergency in Hubei Province in China;5) our insufficiency: □whether there was a significant difference in infectivity between the latent and the infected and it required to be further confirmed in clinical practice; □the blocked and isolated measures had been implemented differently in different areas due to different economic, cultural and management levels; □the maintaining time of the blocked and isolated measures is still in suspense; □whether the epidemic will relapse owing to the surveillance paralysis and the complicated population migration caused by returning to work in a large number of enterprises; □for SEIR model by itself, the limitations on considering little on other vital factors, such as the improvement of prevention, treatment experience and control measures, the advent of new medication, building new hospitals to shorten waiting time in hospital, are not considered, in addition, the setting of each coefficient is still subjective based on clinical practice.

Substantially, COVID-19 is only a local outbreak in Hubei Province, China. It can be probably avoided the pandemic of global SARS-CoV-2 cases rise with the great efforts by Chinese government and its people.

## Data Availability

1)National Health Commission of People’s Republic of China(www.nhc.gov.cn)
2)Tencent's big data(https://heat.qq.com/)and Location Intelligence(www.wayhe.com)
3)Hubei Provincial Center for Disease Control and Prevention (http://wjw.hubei.gov.cn/)

## Contributors

Xiaoping Chen designed the study. Peihua Feng designed the mathematical mode. Xifei Yang collected and analysed data. Wenliang Lu manufactured the table and figures. Gang Wei, Ozarina and Rui Li wrote the manuscript.

## Acknowledgements

We thank Peihua Feng from School of Aerospace Engineering, Xi’an Jiaotong University (Xi’an, China) for technical support.

## References

1 Zhu, N., Zhang, D., Wang, W., et al. A Novel coronavirus from patients with pneumonia in china, 2019. N. Engl. J. Med. 2020, DOI:10.1056/NEJMoa2001017.

2 Imai N, Cori A, Dorigatti I, et al. MRC Centre for Global Infectious Disease Analysis:Wuhan coronavirus reports 1–3. January, 2020. https://www.imperial.ac.uk/mrc-global-infectious-disease-analysis/news--wuhan-coronavirus (accessed Jan 25, 2020).

3 Read, J. M., Bridgen, J. R., Cummings, D. A., et al. Novel coronavirus 2019-nCoV: Early estimation of epidemiological parameters and epidemic predictions. medRxiv 2020, DOI:org/10.1101/2020.01.23.20018549.

4 Joseph T Wu, Kathy Leung, Gabriel M Leung, et al. Nowcasting and Forecasting the Potential Domestic and International Spread of the 2019-nCoV Outbreak Originating in Wuhan, China: A Modelling Study. Lancet. Jan. 31, 2020 DOI: 10.1016/S0140-6736(20)30260-9.

5 Chen, T., Rui, J., Wang, Q., et al. A mathematical model for simulating the transmission of Wuhan novel coronavirus. bioRxiv 2020, DOI: 10.1101/2020.01.19.911669v1.

6 Li Q, Guan X, Wu P, et al. Early transmission dynamics in Wuhan, China, of novel coronavirus-infected pneumonia. N Engl J Med. Jan. 28, 2020, DOI:10.1056/NEJMoa2001316.

7 Cheng, V. C. C., Wong, S. C., et al. Preparedness and proactive infection control measures against the emerging Wuhan coronavirus pneumonia in China. J. Hosp. Infect. 2020, DOI:10.1016/j.jhin.2020.01.010.

8 Liu T, Hu J, Kang M, et al. Transmission dynamics of 2019 novel coronavirus (2019-nCoV). bioRxiv 2020; published online Jan 26, 2020. DOI: http://doi.org/10.1101/2020.01.25.919787 (preprint).

9 Zhao S, Ran J, Musa SS, et al. Preliminary estimation of the basic reproduction number of novel coronavirus (2019-nCoV) in China, from 2019 to 2020: a data–driven analysis in the early phase of the outbreak. bioRxiv 2020; published online Jan 24. DOI:10.1101/2020.01.23.916395 (preprint).

10 Diekmann O, Heesterbeek JAP. Mathematical epidemiology of infectious diseases: model building, analysis and interpretation. Chichester: John Wiley, 2000.

11 Egger, M., Johnson, L., Althaus, C., et al. Developing WHO guidelines: Time to formally include evidence from mathematical modelling studies. F1000Res. 2017, 6, 1584.

12 Rothe, C., Schunk, M., Sothmann, P., et al. Transmission of 2019□nCoV infection from an asymptomatic contact in Germany. N. Engl. J. Med. 2020, DOI:10.1056/NEJMc2001468.

13 Huang, C. Wang, Y., Li, X., et al. Clinical features of patients infected with 2019 novel coronavirus in Wuhan, China. Lancet, 2020, DOI:10.1016/S0140-6736(20)30183-5.

14 Shen, M., Peng, Z., Xiao, Y., et al. Modelling the epidemic trend of the 2019 novel coronavirus outbreak in China. bioRxiv 2020, DOI:10.1101/2020.01.23.916726.

15 Bauch CT, Lloyd-Smith JO, Coffee MP, et al. Dynamically modeling SARS and other newly emerging respiratory illnesses: past, present, and future. Epidemiology. 2005 Nov 1:791–801.

16 Chowell G, Castillo-Chavez C, Fenimore PW, et al. Model parameters and outbreak control for SARS. Emerging Infectious Diseases. 2004 Jul;10 (7):1258.

17 Cohen, J.; Normile, D. New SARS□like virus in China triggers alarm. Science 2020, 367, 234–235.

18 Lessler J, Reich NG, Brookmeyer R, et al. Incubation periods of acute respiratory viral infections: a systematic review. Lancet Infect Dis. 2009;9 (5):291–300. DOI:10.1016/S1473-3099(09)70069-6.

19 Lloyd-Smith JO, Schreiber SJ, Kopp PE, et al. Superspreading and the effect of individual variation on disease emergence. Nature 2005; 438: 355–59.

20 Lipsitch M, Cohen T, Cooper B, et al. Transmission dynamics and control of severe acute respiratory syndrome. Science 2003; 300: 1966–70.

21 Wu, P., Hao, X., Lau, E. H., et al. Real-time tentative assessment of the epidemiological characteristics of novel coronavirus infections in Wuhan, China, as at 22 January 2020. Eurosurveillance 2020, 25, DOI:10.2807/1560-7917.ES.2020.25.3.2000044.

22 Biao Tang, Xia Wang, Qian Li. et al. Estimation of the Transmission Risk of the 2019-nCoV and Its Implication for Public Health Interventions. J Clin Med, 9 (2), Feb. 7, 2020. DOI: 10.3390/jcm9020462.

23 Riley S, Fraser C, Donnelly CA, et al. Transmission dynamics of the etiological agent of SARS in Hong Kong: impact of public health interventions. Science 2003; 300: 1961–66.

24 The 2019-nCoV Outbreak Joint Field Epidemiology Investigation Team, Li Q. An outbreak of NCIP (2019-nCoV) infection in China—Wuhan, Hubei Province, 2019–2020. China CDC Weekly 2020;2: 79–80.

25 Bootsma MC, Ferguson NM. The effect of public health measures on the 1918 influenza pandemic in U. S. cities. Proc Natl Acad Sci USA 2007; 104: 7588–93.

26 Hui, D. S., Azhar, E. E. I., Madani, T. A., et al. The continuing 2019□nCoV epidemic threat of novel coronaviruses to global health□The latest 2019 novel coronavirus outbreak in Wuhan, China. Int. J. Infect. Dis. 2020, 91, 264–266.

27 Bogoch II, Watts A, Thomas-Bachli A, et al. Pneumonia of Unknown Etiology in Wuhan, China: Potential for International Spread Via Commercial Air Travel. Journal of Travel Medicine. Jan. 14, 2020. DOI: 10.1093/jtm/taaa008.

